# Mitigating norovirus spread on cruise ships: A model-based assessment of diagnostic timing and isolation

**DOI:** 10.1101/2024.12.09.24318708

**Authors:** Alfredo De Bellis, Andrea Bizzotto, Lemonia Anagnostopoulou, Leonidas Kourentis, Valentina Marziano, Varvara Mouchtouri, Stefano Merler, Giorgio Guzzetta

## Abstract

**Background:** Acute gastroenteritis outbreaks caused by noroviruses are a common public health issue on cruise ships. Understanding the main drivers of sustained outbreaks is critical for evaluating the effectiveness of preventive interventions such as the isolation of infected individuals.

**Methods:** We analyzed a line-list of 121 cases from a norovirus outbreak on a cruise calling Mediterranean ports (cumulative incidence among passengers 9.7%). We used a Bayesian inference model to reconstruct likely transmission chains, taking into account different transmission settings and the isolation of cases after diagnosis. We then calibrated a branching process model to simulate alternative isolation scenarios and estimate their effectiveness in reducing transmission.

**Results:** Reconstructed transmission chains revealed a high heterogeneity in individual transmission, with 57% (95% CrI: 48%-65%) of secondary cases caused by 10% of infected individuals (here termed “superspreaders”). Superspreaders exhibited longer diagnostic delays (mean 83 hours, 95% CrI: 70-96 hours) compared to other infectors (mean 47 hours, 95% CrI: 44-50 hours) and a halved frequency of vomiting and diarrhea episodes. The 72-hour isolation protocol implemented during the outbreak averted 71% of potential cases compared to a no-intervention scenario, halving the effective reproduction number from 9.8 (95%CrI of the mean: 7.1-12.7) to 4.9 (95%CrI: 3.0-7.1). Reducing diagnostic delays further reduced the effective reproduction number, resulting in lower case numbers and probability of sustained outbreaks.

**Conclusions:** Timely diagnosis and isolation have a remarkable impact on norovirus containment on cruise ship outbreaks. Targeted information campaigns encouraging passengers to seek immediate medical assistance upon gastrointestinal symptoms can significantly improve outbreak management.

## Background

Norovirus is a main cause of outbreaks of gastroenteric infections (GI) on cruise ships [1]. Norovirus can spread explosively on board, either through contaminated food, water or surfaces, or through person-to-person transmission. In turn, person-to-person transmission can happen directly during close social contacts, or via the ingestion of virus particles aerosolised through vomitus or feces [2].

In addition to direct consequences on travellers’ health, outbreaks of norovirus may cause the disruption of holidays for affected passengers and co-travellers, temporary shortages of crew members for routine cruise operations, and high costs related to the implementation of infection control measures for cruise companies, with a remarkable economic burden [3]. If we exclude the temporary effect of the COVID-19 pandemic on the cruise passenger market [4], the number of passengers worldwide has seen a steady and rapid expansion since 2004 [5], suggesting a potentially higher future impact of norovirus outbreaks on the tourism industry in the near future.

Standards for hygiene and plans for prevention, mitigation and management measures (PMM) have been defined by multiple international institutions [6-8]. Pre-embarkation screening, syndromic surveillance on board, isolation of infected individuals, application of environmental disinfection and education of crew and passengers on hand washing and rapid reporting of symptoms have been identified among the main preventative measures [6-8]. Outbreak management measures may include active surveillance, enhanced disinfection of public surfaces, discontinuation of self-service restaurants, and social distancing [6-8].

A better understanding of the dynamics of norovirus spread on board is critical to assess the effectiveness of control interventions. In this work, we analyze an individual line-list of GI cases from a large outbreak on a cruise ship [10] to probabilistically reconstruct transmission chains on board, assess the importance of diagnostic delays on transmission and evaluate the effectiveness of alternative case isolation scenarios.

## Materials and Methods

We considered data from an outbreak of 121 GI cases, reported over a seven days voyage on a cruise ship calling Mediterranean ports [10]. Data consisted in a line-list of reported cases with information on case gender, role (passenger or crew member), cabin number, time of symptom onset, date of diagnosis, type of symptoms (vomiting or diarrhea) and number of vomiting and diarrhea episodes until diagnosis. For 65 of 121 cases there was a PCR confirmation of norovirus presence in stools, therefore we assumed that all 121 GI cases were caused by norovirus. The cruise carried 1229 passengers and 487 crew members (9.7% attack rate among passengers, 0.4% among crew members, 7.1% overall). Summary information for the analyzed outbreak is reported in Table 1, and the number of cases by time of symptom onset (panel A) and by date of diagnosis (panel B) are shown in Figure 1. Data processing was performed in R, version 4.4.0 (2024-04-24).

**Table 1.** Population and symptom characteristics for the considered outbreak data.

| Variable | Number of cases (%, N=121) |
| --- | --- |
| <u>Symptoms:</u> |  |
| Only vomit | 1 (0.8) |
| Only diarrhea | 25 (20.7) |
| Both | 95 (78.5) |
| <u>Gender:</u> |  |
| Male | 54 (44.6) |
| Female | 67 (55.4) |
| <u>Role:</u> |  |
| Passengers | 119 (98.3) |
| Crew members | 2 (1.7) |

**Figure 1.**
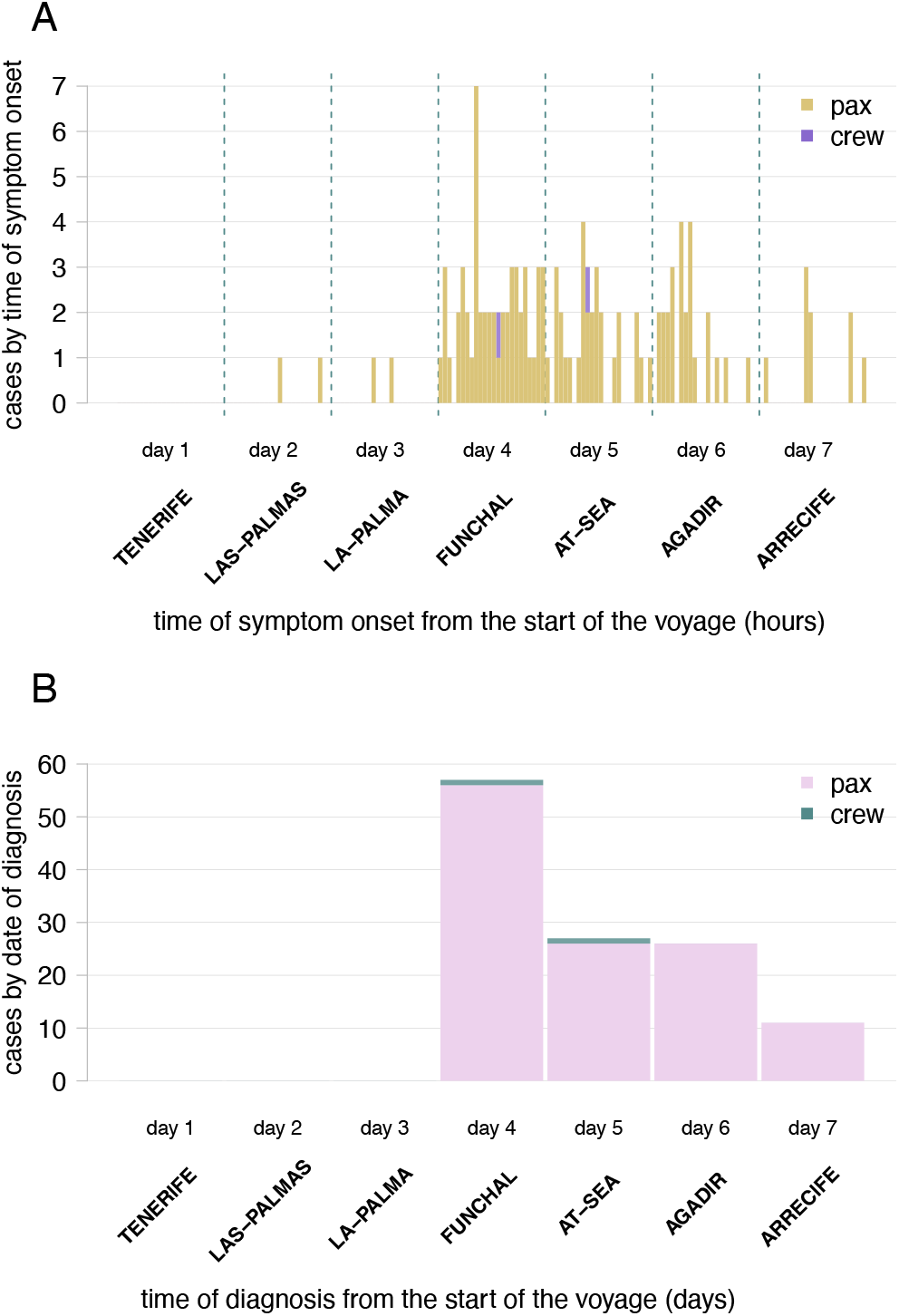
Epidemic curves, disaggregated by passengers (pax) and crew members (crew). **A**. Hourly time series by symtpom onset. **B**. Daily time series by date of diagnosis.

We applied a Bayesian model to probabilistically reconstruct transmission chains (i.e., who infected whom) [11,12,13]. For each susceptible individual on board, the model defines a force of infection (FOI) exerted over time by infected individuals, considering the possibility of cases infected before first embarkation, of infection among people sharing cabins during night-time, transmission among travellers on board of the ship, and acquisition of infection during visits in ports of calling. We implemented the model in C, using the gcc compiler and the GSL (GNU Scientific Library) for external dependencies.Specifically, at any time t, we assume that each susceptible individual j is exposed to a FOI *λ*_*j*_:

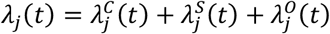

Where:

- 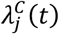 represents the FOI from infected cabinmates during the night;
- 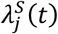 represents the FOI from infected individuals onboard the ship in public spaces;
- 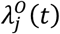 represents the FOI from infected individuals offboard during visits at ports.

A mathematical formulation of the different contributions to the FOI is reported in the Appendix. Briefly, the FOIs depend on the distribution of the generation time, modeled as a Gamma distribution *Γ* with a mean of 87.6 hours [14], and includes increased transmission rates for cases who reported vomiting (*ρ*^*V*^ = 2.12-fold increase), diarrhea (*ρ*^*D*^ = 1.39-fold), or both (*ρ*^*V*^ · *ρ*^*D*^ = 2.95-fold) [15]. The model explicitly represents the isolation protocol observed on board, by assuming that diagnosed individuals are immediately confined in their cabins for 72 hours. Isolation results in the impossibility to transmit to people other than cabin members. A full mathematical specification of the FOI is provided in the Appendix. The model assigns likely infectors by selecting which potential index case contributed the most to an individual’s FOI at their time of infection. We implemented the model in C, using the gcc compiler and the GSL (GNU Scientific Library) for external dependencies.

The model was calibrated using a Markov Chain Monte Carlo (MCMC) procedure with Metropolis-Hastings sampling algorithm; free model parameters were the transmission rates for each route of infection (*β*_*c*_, *β*_*s*_, *β*_*P*_), the average prevalence of norovirus in port communities (*α*), the increase in transmissibility in public areas among cabinmates (*ϕ*) and the unknown infection times (*τ*_*j*_) of each case *j*.

The overall likelihood used within the MCMC is given by:

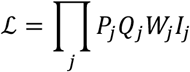

where *P*_*j*_ represents the likelihood that individual *j* was infected by an infector *k*_*j*_:

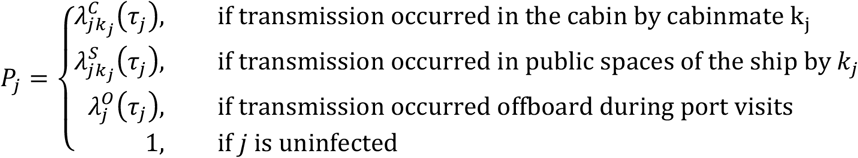

*Q*_*j*_ represents the likelihood that j was not infected (until τ_*j*_ or until the end of the cruise):

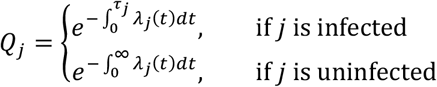

*W*_*j*_ is the likelihood of the incubation period estimated for individual *j*:

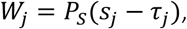

where *P*_*S*_ is the distribution of the incubation period, assumed to be a log-normal with mean 32.6 hours [16] and *s*_*j*_ is the time of symptom onset of *j*.

Finally, *I*_*j*_ is the likelihood of having been infected before embarkation (imported):

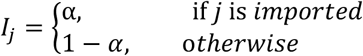

Further details on the definition of the likelihood and MCMC procedure can be found in the Appendix.

Since the data reported the date but not the time of diagnosis for each case, model calibration was repeated Z=50 times after imputing for all reported cases their time of diagnosis (in hours). The time was sampled uniformly over the date of diagnosis, excluding night hours (between midnight and 8am) and times preceding the time of symptom onset if this occurred on the same date as diagnosis. M=2500 parameter values and reconstructed transmission chains were sampled from the posterior distributions of the Z calibration procedures and pooled together to obtain the final results. To evaluate results with respect to the potential underreporting of cases, we run sensitivity analyses where we re-calibrated the model assuming levels of underreporting of 23% and 40% [17-18]. A description of the main model parameters is provided in Table 2. Full model specifications are provided in the Appendix.

**Table 2.** Model parameters. For calibrated values, the mean (95% CrI) of the posterior distribution is provided. For distributions, the mean value is provided (the full specifications of the distribution are reported in the Appendix).

| Parameter | Description | Value | Reference |
| --- | --- | --- | --- |
| Common parameters |  |  |  |
| $\Gamma$ | Generation time distribution (Gamma) | mean: 87.6 hours | [14] |
| $P_S$ | Incubation period distribution (log-normal) | mean: 32.6 hours | [16] |
| Transmission chain reconstruction model |  |  |  |
| $\rho^V$ | Relative transmissibility for infectors with vomiting | 2.12 | [15] |
| $\rho^D$ | Relative transmissibility for infectors with diarrhea | 1.39 | [15] |
| $\beta_C$ | Scaling factor for transmissibility inside cabins | 1.56 (0.04-0.35) | Calibrated |
| $\beta_S$ | Scaling factor for transmissibility in public areas of the ship | 8.1 (5.1-22.5)·10<sup>-3</sup> | Calibrated |
| $\beta_P$ | Scaling factor for transmissibility offboard during port visits | 2.03 (0.04-6.65)·10<sup>-2</sup> | Calibrated |
| $\alpha$ | Relative prevalence in port population | 0.36 (0.05-0.96)% | Calibrated |
| $\phi$ | Relative transmissibility for cabin-mates infectors on ship | 1.23 (1.01-1.52) | Calibrated |
| Branching process model |  |  |  |
| $\Delta$ | Distribution of diagnostic delay | mean: 17.0 hours | Empirical |
| $R_0$ | Basic reproduction number | 15.6 (11.5-20.4) | Calibrated |
| $\nu$ | Overdispersion of the offspring distribution | 0.13 (0.10-0.18) | Calibrated |

To assess the overall effectiveness of alternative isolation protocols and the impact of diagnostic delays, we additionally developed a branching process model [19], where each infected individual is assumed to generate a theoretical number of cases sampled from a negative binomial distribution with mean proportional to the basic reproduction number ***R***_0_ and overdispersion *ν*. The time of theoretical secondary infections was distributed according to the distribution of the generation time [14]; infections whose sampled time would fall within the isolation period of the infector, or after the end of the cruise, were discarded from the realized infections. The branching process model does not explicitly distinguish cabin transmission or importation of cases during tours.

We assumed that two cases were infected before embarkation (based on results from the reconstructed transmission chains), and their symptom onset time was fixed at those of the first two symptomatic cases in the dataset; their time of infection was established by subtracting an incubation period sampled from the same log-normal distribution used for the transmission chain reconstruction model [16], until the assigned infection time preceded embarkation. For each case transmitted on board, the time of symptom onset was assigned by adding to the time of infection an incubation period sampled from the same distribution. The time of diagnosis (corresponding to the time of isolation start) was assigned for all infections by adding a diagnostic delay sampled from an empirical distribution Δ determined from observed outbreak data; infections for which the assigned diagnosis time fell after the end of the cruise were considered as unreported. The time at which isolation ended for each case was assigned by adding a fixed duration of isolation (72 hours in the baseline analysis) to the time of diagnosis.

The observed outbreak may be interpreted as an individual realization of a stochastic process, which the branching process aims to represent. Therefore, we calibrated free model parameters (***R***_3_ and ν) by using a particle filtering approach [20] weighing model trajectories (“particles”) by the mean squared error (MSE) between the modelled and observed epidemic curves. The MSE for each particle *w* was computed as:

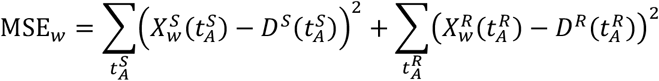

where 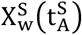 is the time series of cases by time of symptom onset for particle 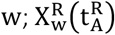 is the time series by date of diagnosis for the particle 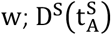 is the time series of observed cases by time of symptom onset; 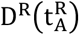 is the time series of observed cases by date of diagnosis. The variables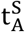 and 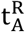 represent the aggregated times of analysis (i.e., 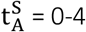 hours, 4-8 hours, 8-12 hours, etc., for time series by time of symptom onset and 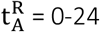 hours, 24-48 hours, 48-72 hours, etc., for time series by date of diagnosis). Parameters were explored by grid search (see Appendix for details).

The best-fitting parameter sets were then used to simulate seven alternative intervention scenarios. In the first four, isolation of diagnosed cases was reduced from 72 hours (baseline scenario, i.e. the strategy actually implemented on board) to 48 and 24 hours or was not done altogether. In two other scenarios we maintained the 72 hours isolation, but the empirical diagnostic delays after symptoms were either increased or reduced by 50%. Finally, we considered a “perfect” isolation scenario, where for all cases isolation was immediate after the development of first symptoms and lasted until the end of the cruise. Model outputs were the relative reduction in the total number of cases observed in the cruise compared to a scenario with no interventions, and the effective reproduction number, measured as the mean number of secondary cases realized in the cruise by infections occurring within day 2 of the voyage. Considering only infections from the early days of voyage reduces the right-censoring effects due to infections that would occur after the end of the cruise. Full details on the branching process model and calibration are reported in the Appendix.

## Results

In the model-reconstructed transmission chains for the considered outbreak, 4.5% (95% credible interval, CrI: 1.7%-8.3%) of infections on average occurred before embarkation, 7.3% (95%CrI: 2.5%-10.7%) were transmitted in the cabin and 88.0% (95%CrI: 82.7%-93.4%) occurred in public spaces of the ship. Acquisition during visits at ports was estimated to be negligible.

Based on the reconstructed transmission chains, the effective reproduction number (i.e., the number of secondary cases caused by a single infectious individual) declined over time for cases infected later, together with their delay between the time of infection and that of diagnosis, hereafter termed “infection diagnostic delay” (Figure 2). Cases already carrying the virus before embarkation on the cruise ship caused an average of about 7.6 secondary infections each (95%CrI of 4.4-14.5). Individuals infected during the first 24 hours of cruise went on to cause about 3.0 (95%CrI 1.5-5.2) secondary cases each; the effective reproduction number decreased briskly in the following days, and crossed the epidemic threshold of 1 for infections that occurred at after 24 hours since the start of the cruise.

**Figure 2.**
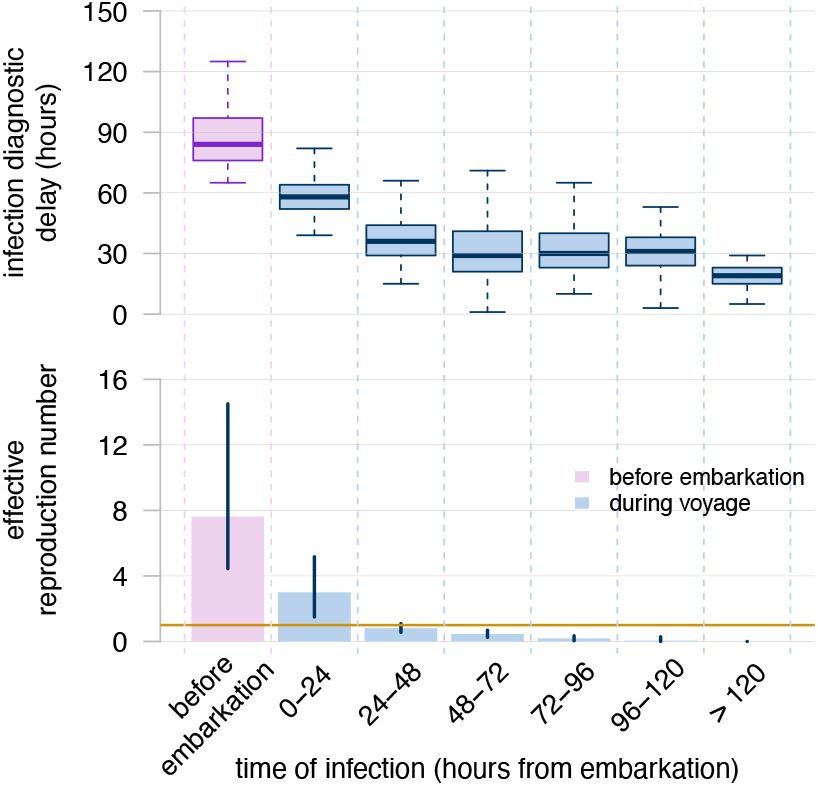
Reproduction number and infection diagnostic delay over the course of the outbreak. Boxplots (**top**) show the distribution of infection diagnostic delays (in hours) for cases infected on each day of the voyage. The bar plot (**bottom**) represents the estimated effective reproduction number for cases infected on each day of the voyage. The bars indicates the mean estimate, while the error bars indicate the 95%CrI of the mean over all the reconstructed chains

The model estimates that 60% (95%CrI: 55%-65%) of cases did not transmit the infection further, while 11% of individuals went on to cause 3 or more secondary cases (Figure 3A). Fitting negative binomial distributions to the offspring distributions associated to model-reconstructed transmission chains resulted in a mean overdispersion parameter of 0.42 (95%CrI: 0.28-0.62), indicating the existence of superspreading individuals disproportionately contributing to transmission. This result is confirmed in Figure 3B, displaying the cumulative proportion of cases caused by infectors ranked by their number of secondary cases; the figure shows that the top 20% of infectors were responsible for 77% (95%CrI: 71%-83%) of all cases, in accordance with the Pareto rule [21], and the top 10% was responsible for 57% (95%CrI: 48%-65%) of all cases.

**Figure 3.**
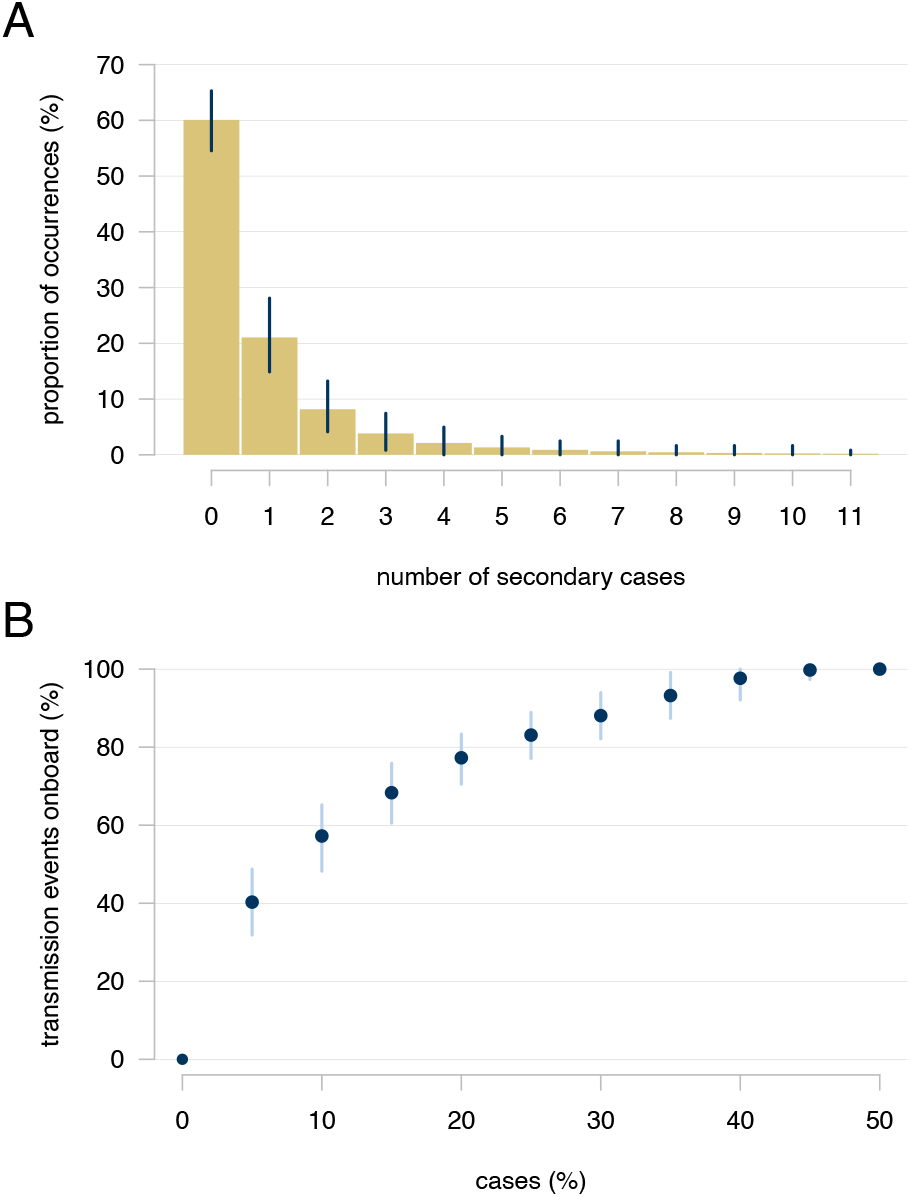
Superspreading in the considered outbreak. **A**. Distribution of the number of secondary cases generated by infected individuals. Bars indicate mean values over 125,000 reconstructed transmission chains, while error bars indicate the 95%CrI; **B**. Cumulative proportion of secondary cases ranked by infectors. The points indicates the average value, while the error bars indicate the 95%CrI over all the reconstructed chains.

Figure 4 shows a disaggregation of the model-estimated mean number of secondary cases by presence of vomit among symptoms, and by infection diagnostic delay. The mean number of secondary cases generated by cases who experienced vomiting episodes was estimated by the model at 1.02 (95%CrI: 0.88-1.15), significantly higher than the corresponding value for cases with no vomiting (mean 0.70, 95%CrI: 0.28-1.28; Student’s t-test p-value << 10^-6^) (Figure 4A). The estimated mean number of secondary cases disaggregated by infection diagnostic delay increased from 0.39 (95%CrI: 0.27-0.51) for infections diagnosed within 2 days, to 1.89 (95%CrI: 1.13-2.81) for delays between 2 and 3 days, and to 5.13 (95%CrI: 3.14-8.25) for delays larger than 3 days (Figure 4B).

**Figure 4.**
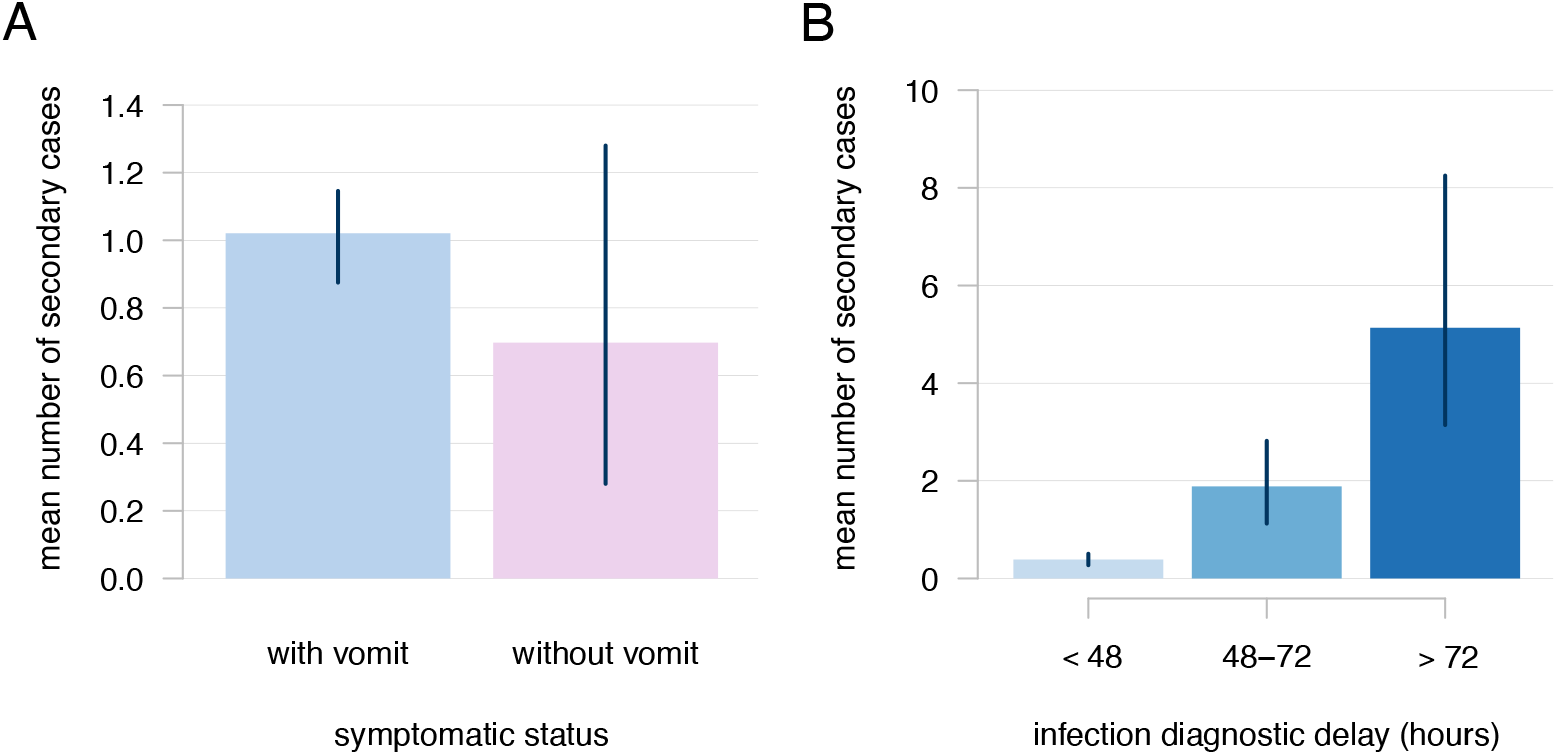
**A** Number of secondary cases generated by individuals with and without vomiting episodes; **B** Number of secondary cases stratified by infection diagnostic delay. Bars indicates mean values, error bars indicate the 95%CrI of the mean over all the reconstructed chains.

The model estimated a mean number of secondary cases for the top 10% of infectors of 5.64 (95%CrI: 4.83-6.42), 12.5-fold higher than the remainder of cases (0.45; 95%CrI: 0.38-0.54) (Figure 5A). The mean diagnostic delay for the top 10% of infectors was 83 hours (95%CrI: 70-96 hours), much larger than the one for the remainder of cases (47 hours, 95%CrI: 44-50 hours) (Figure 5B). The top 10% of infectors experienced gastrointestinal symptoms (vomiting or diarrhea) with a halved frequency (mean 4.8, 95%CrI: 3.5-6.4 per day) compared to the remainder of infected individuals (mean 9.8, 95%CrI: 9.1-10.6 per day) (Figure 5C).

**Figure 5.**
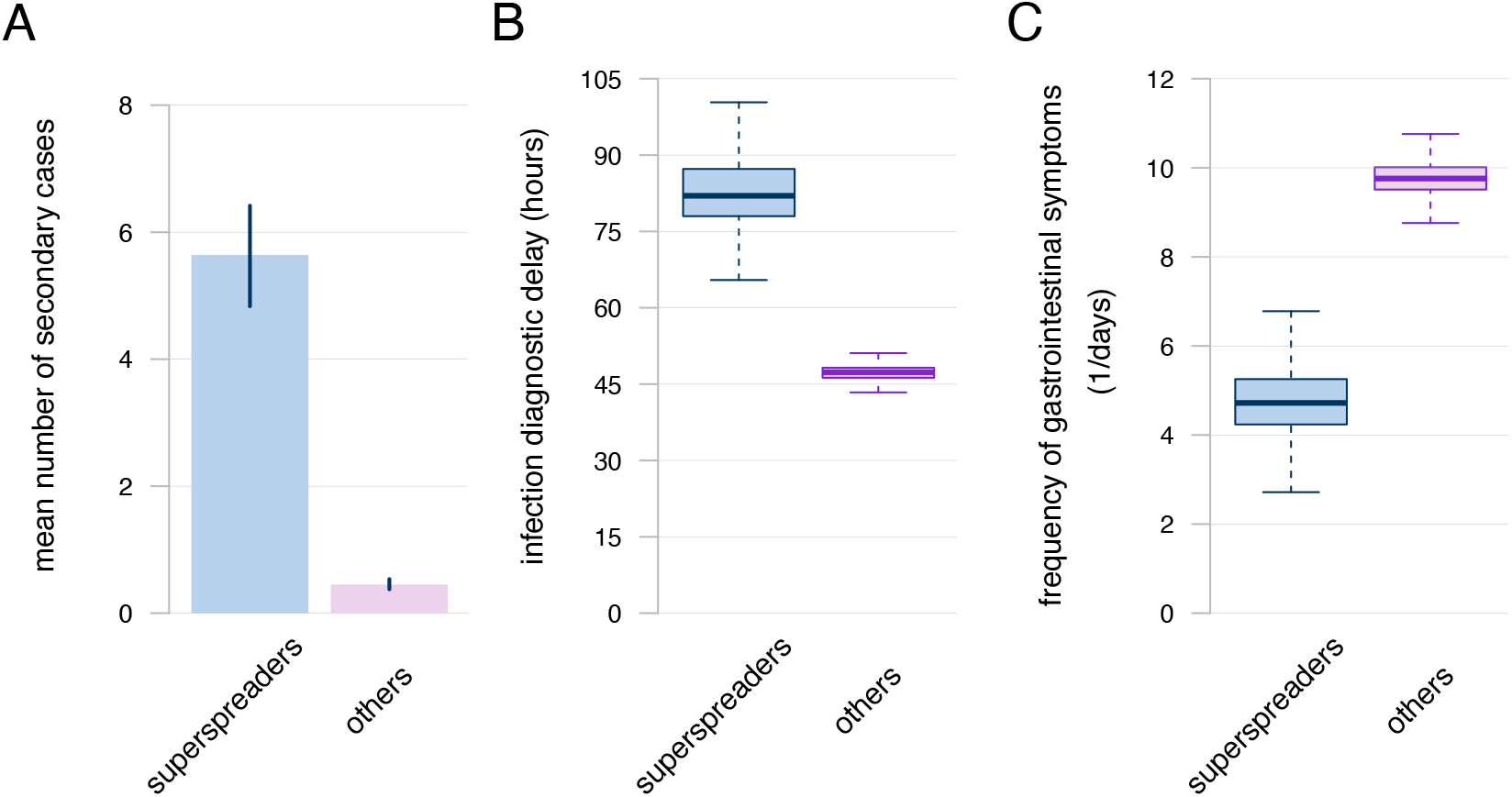
Characteristics of superspreaders (top 10% of infectors) compared to other cases. **A** Mean number of secondary cases. Bars indicate the mean estimate, error bars indicate the 95%CrI across the 125000 reconstructed chains. **B** Infection diagnostic delays. **C** Frequency of gastrointestinal symptoms per day. Boxplots represent the mean, IQR and 95%CrI of the mean across the 125000 reconstructed chains.

All findings remained robust when considering a potential underreporting of cases up to 40% (see Appendix).

The calibration of the branching process model to outbreak data resulted in an estimated value for the basic reproduction number ***R***_0_ of 15.6 (95%CrI 11.5-20.4) and for the overdispersion in the offspring distribution *ν* of 0.13 (95%CrI 0.10-0.18) (see Appendix). We found that, even in absence of any control strategies, the effective reproduction number on board decreased to 9.8 (95%CrI of the mean: 7.1-12.7) due to the short duration of the cruise, since a fraction of secondary cases would be transmitted by infectious individuals after disembarkation (Figure 6A). The isolation of cases in their cabins for 24, 48 and 72 hours further reduced the reproduction number to 7.8 (95%CrI: 5.3-10.5), 6.0 (95%CrI: 4.2-7.9) and 4.9 (95%CrI: 3.0-7.1) respectively. Reductions and increases by 50% in observed diagnostic delays after symptoms (maintaining isolation for 72 hours as in the baseline) would have a mild impact on the effective reproduction number. Even in a perfect control scenario where all cases are diagnosed and isolated instantaneously after symptom onset and until the end of the cruise, the reproduction number would still be significantly above the epidemic threshold at 3.8 (95%CrI of the mean: 2.1-5.7). Perfect isolation would have led to averting on average 89% of cases that would occur in absence of isolation (Figure 6B). We estimate that the implemented control scenario with 72-hours isolation of a case after diagnosis averted about 71% of the potential cases. The proportion of averted cases goes down to 60% in the case of 72-hours isolation but with 50% longer symptom diagnostic delays, and up to 81% with 50% shorter symptom diagnostic delays. Reducing the isolation duration to 48 and 24 hours respectively would avoid 67% and 50% of cases compared to a no-intervention scenarios. The probability of observing an outbreak, which is defined as a cruise where the number of total cases exceeds 2% of the passenger population, was above 90% in all scenarios, except those envisioning a marked reduction of diagnostic delays.

**Figure 6.**
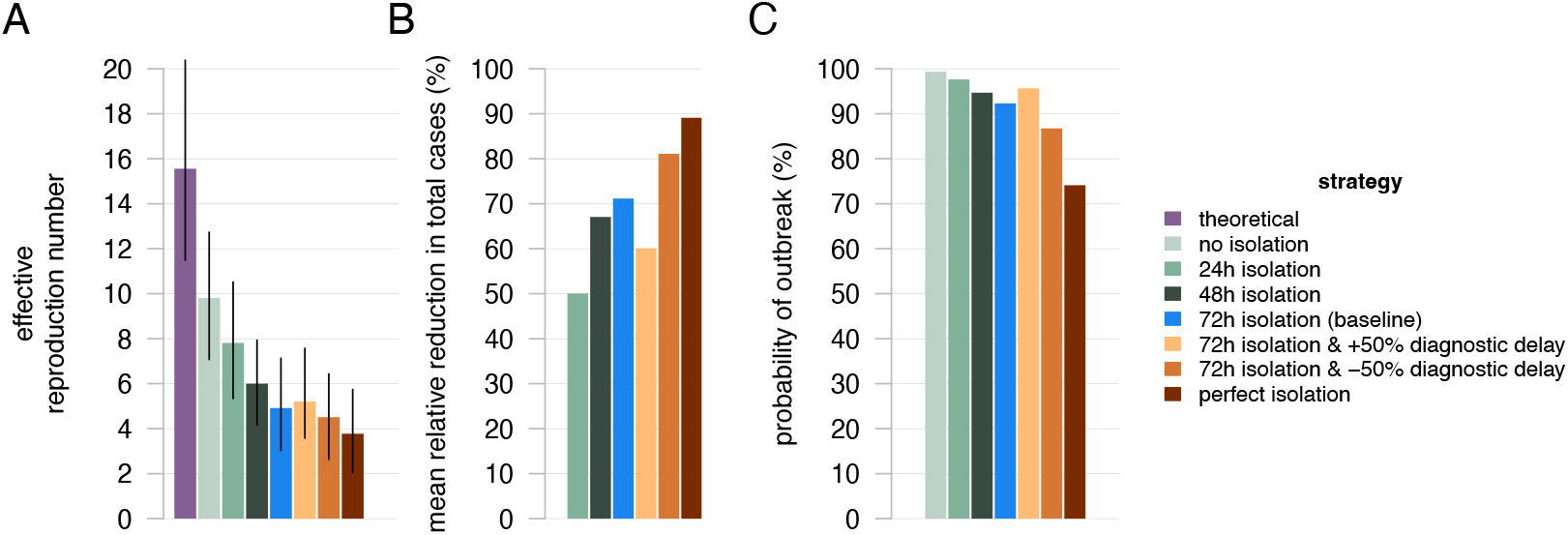
Impact of alternative control scenarios. **A** Effective reproduction number. For the theoretical basic reproduction number, the bar represent the mean and error bar represent the 95%CrI of the posterior distribution estimated via MCMC. For scenarios, bars indicate mean values over 100 simulations and error bars indicate the 95%CrI of the mean. **B** Mean relative reduction in the total expected cases compared to a scenario with no interventions. **C** Probability of an outbreak (defined as >2% of passengers infected).

## Discussion

We reconstructed likely transmission chains during a large norovirus outbreak on a cruise ship calling Mediterranean ports, providing insights in the transmission patterns on board.

We estimated that the majority of cases occurred in public spaces on the cruise ship (almost 90%), with limited transmission in cabins and a negligible number of infections acquired during tours in visited ports. We identified a strong degree of superspreading, with 20% of patients being responsible for almost 80% of the infections diagnosed on board. Delays between infection and diagnosis emerged as a key driver of outbreak dynamics. The association of longer diagnostic delays with higher numbers of secondary cases corresponds to the increased opportunities to transmit the infection to others through social contacts on the ship before being isolated in their cabins for 72 hours. The top 10% of infectors, who transmitted on average 12.5 times more cases than the rest of the infected individuals, were characterized by diagnostic delays that were on average twice the ones for the rest of cases. The halved frequency of gastrointestinal symptoms in the top 10% of infectors compared to the rest of cases may explain why these individuals had longer delays in health-seeking behavior, indicating that the difficulty in timely identifying mild cases may play a critical role in transmission dynamics. Awareness of the existence of norovirus spread on board after the first diagnosed cases, diffused among passengers via information campaigns on board and word-of-mouth, may have led to the reduction over time of diagnostic delays and timely isolatioon of cases, facilitating outbreak management. Results were robust when considering the possibility of a non-negligible fracton of infections remaining underreported at the end of the cruise [17, 18].

Simulations based on a branching process model estimated the basic reproduction of norovirus on board of the cruise at about 15, a value that is compatible with high values estimated in other crowded settings [9, 14]. This theoretical value does not correspond to the actual average number of secondary infections (“effective reproduction number”) on board because a proportion of all infections would occur after the end of voyage duration, which was 7 days in the considered cruise. Longer voyages are likely subject to higher effective reproduction numbers. The implemented protocol of isolation for 72 hours after diagnosis reduced the estimated effective reproduction number to 4.9 (95%CrI 3.0-7.1). This value is consistent with the 95%CrI of 4.4-14.5 for individuals who carried the virus at embarkation and of 1.5-5.2 for individuals infected on the first day, estimated by the transmission chain reconstruction model.

We estimated that the implemented isolation protocol has avoided about 71% of potential cases that would be observed in its absence. Choosing protocols with shorter durations of isolation would have avoided significantly less cases, while halving diagnostic delays after symptoms might further improve the effectiveness of the protocol, increasing the proportion of avoided cases to 80%. Even in absence of diagnostic delays and with permanent isolation of cases, the reproduction number would still remain largely above the epidemic threshold of 1 and a >70% probability of observing a cumulative attack rate among passengers higher than 2%. Previous modeling studies [9] have estimated the effectiveness of isolation in reducing the outbreak size, but did not consider the effect of diagnostic delays, assuming sudden isolation of cases.

This study has several limitations. First, we did not have sufficient data to explicitly model potential environmental or foodborne transmission. Although the literature suggests direct person-to-person transmission is predominant [9], environmental contamination could also play a role in norovirus outbreaks and requires further investigation. However, a recent systematic review and meta-analysis found that person-to-person was the most frequent mode of transmission in 35 out of 45 outbreaks considered [22]. Second, the present analysis is based on a single outbreak, albeit characterized by a large cumulative incidence (9.7% among passengers), and caution should be exercised when generalizing the findings to other settings or outbreaks. The unique conditions of a cruise ship may not fully represent other environments where norovirus spreads. Our finding that the risk of importation from port visits is negligible may not hold for other geographical contexts with higher endemic norovirus prevalence and less strict hand hygiene and food handling norms. Given the small number of cases occurring in crew, we approximated transmission as occurring entirely in the passenger population and we could not study the effect of isolation separately on the passenger and crew populations.

Our findings confirm the difficulty of halting norovirus transmission on board, and the fundamental role of timely isolation of cases to control transmission in confined settings like cruise ships. The low frequency of symptoms and long diagnostic delays in superspreaders suggest the importance of passenger education towards seeking immediate health assistance when experiencing gastrointestinal symptoms on board, to reduce the probability of outbreaks and their cumulative incidence.

## Data Availability

All data produced in the present study are available upon reasonable request to the authors

## APPENDIX

### 1. Inference of transmission pairs

#### 1.1 Model description

At any time t, we assume that each susceptible individual j is exposed to a force of infection (FOI):

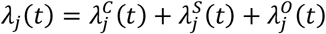

Where:

- 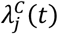 represents the FOI from infected cabinmates,
- 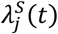 represents the FOI from infected individuals onboard the ship outside cabin (in public spaces)
- 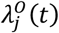 represents the FOI from infected individuals offboard during visits at ports.

The FOI from infected cabinmates is defined as:

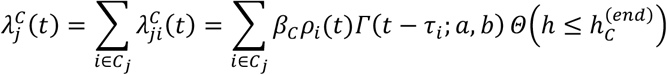

where:

- *C*_*j*_ is the set of individuals who share the same cabin as individual j;
- *β*_*C*_ is a free model parameter scaling the transmissibility of norovirus inside cabins;
- *ρ*_*i*_(*t*) is the relative increase in infectivity of the potential infector *i* based on their symptoms status at time *t*: 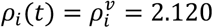 if the individual *i* had at least one vomit episode, 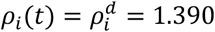 if the individual *i* had at least one diarrhea episode, 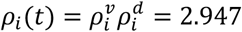 if the individual *i* experienced both kinds of symptomatic episodes [S1], *ρ*_*i*_(*t*) = 1 if the individual *i* did not have symptoms at time *t*;
- *Γ*(*x; a, b*) is the discretized distribution of the generation time, i.e. the time that elapses between infection episodes of an infector-infectee pair, assumed to be distributed as a Gamma with shape *a* = 3.35 and rate *b* = 0.92 (mean generation time: 3.65 days) [S2];
- *τ*_*i*_ represents the time of infection of individual *i*;
- *Θ*(*X*) is a Heaviside step function that is 1 when the condition *X* is true, and 0 otherwise. It is devised in such a way that the FOI within the cabin occurs only between midnight and 08:00 of each day of navigation, i.e. when passengers are assumed to withdraw in their cabins for sleeping (see Figure S0);
- *h* represents the hour of the day and is calculated by using the modulo operation on the calendar time *t* since the start of the cruise, with respect to a 24-hour cycle. In other words, *h* = *t*_*mod*24_. For example, if *t* = 26 hours then *h* = 26 _*mod*24_= 2, corresponding to 02:00 in the morning. Time *t* is initialized at *t* = 0, which corresponds to the beginning of the first day of travel at midnight (00:00);
- 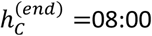 is the hour at which we assume passengers to leave the cabin.

The FOI in public spaces accounts for the contribution from infected individuals (both cabinmates and others) during daytime. This FOI is only applicable to a potential infector-infectee pair if neither individual is in quarantine at the time being considered and at least one of the following condition holds: (i) it is a day of navigation at sea, i.e. without a stop in a port; (ii) the time of the day falls before or after the times of disembarkment for port visits; (iii) both the infector and the infected stay on board of the ship despite the possibility of disembarking to visit the port destination.The FOI from public spaces of the ship is therefore defined as:

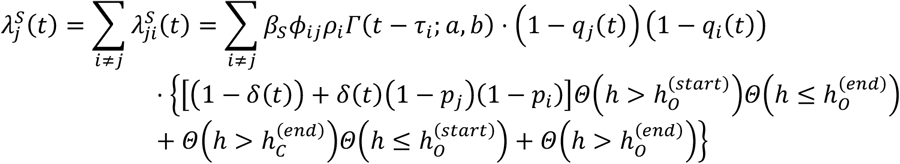

where:

- *β*_*s*_ is a free parameter scaling the transmissibility of norovirus onboard in public spaces of the ship;
- 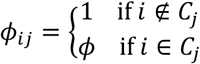 and *ϕ* ≥ 1 is a free parameter accounting for the possibly increased relative transmissibility in public areas among cabinmates, given that they are more likely to spend time in close contact;
- *q*_*j*_(*t*) is a Kronecker delta-function accounting for quarantine of individual *j* over time: it is one if individual *j* is quarantined in cabin at time *t* and zero otherwise.
- *δ*(*t*) is delta-function that equals zero if the day associated to time *t* is a day of navigation at sea, and one otherwise;
- *p*_*j*_ is the probability that the individual *j* goes on the daily visit at ports. We assume that *p*_*j*_ = 90%, if *j* is a passenger and *p*_*j*_ = 0% if *j* is a crew member (i.e., the proportion of crew members disembarking for visiting ports is considered negligible);
- 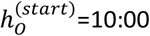 and 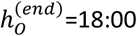 are the hours at which we assume the port visits start and end, respectively;
- *Θ*(*X*) is devised in such a way that the FOI in public spaces occurs only between 08:00 and 10:00 or between 18:00 and midnight, when passengers are assumed to be outside cabins but on the ship, during days of port visits (see Figure S0). During days of navigation at sea, *Θ*(*X*) allows the FOI in public spaces to occur also between 10:00 and 18:00;

The FOI from the general port population accounts for the contribution from infected individuals encountered offboard during the designated port visit hours. This FOI is only relevant if the individual *j* visits a port and is not in quarantine, and is defined as:

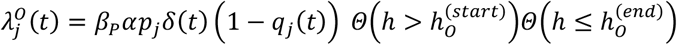

where *α* is a free parameter representing the relative prevalence of norovirus in the general port population and *β*_*P*_ is a free parameter scaling the transmissibility offboard during port visits. *Θ*(*X*) is devised in such a way that the FOI during visits occurs only between 10:00 and 18:00 (see Figure S0).

A schematics of a typical day schedule for a traveler disembarking for a port visit is illustrated in Figure S0.

**Figure S0.**
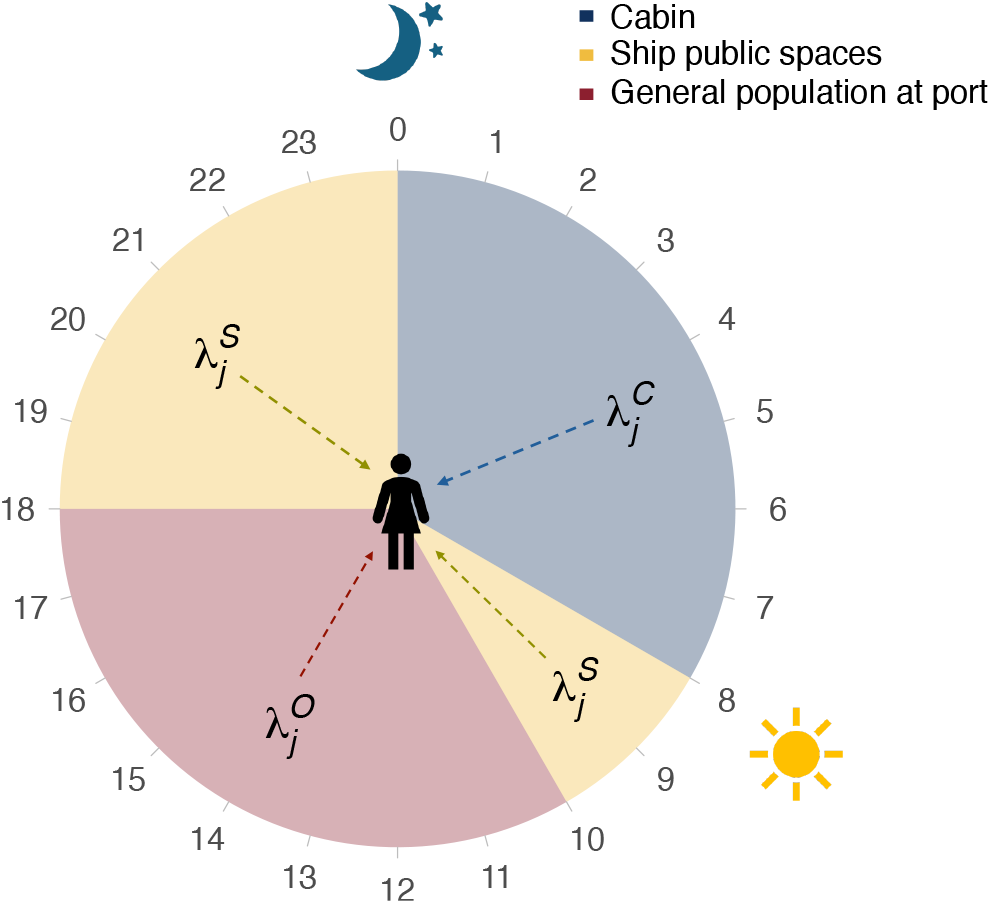
Example of daily schedule and corresponding exposure to different forces of infection for a traveller disembarking for a port visit.

The model assigns a source of infection *k*_*j*_ for all cases by choosing from sources of one of the three settings of transmission (an infector within cabins, an infector in public spaces onboard, or a generic source of infection in offboard port visits) with probability π_*ji*_ proportional to the contribution of each source to the total FOI at the time *τ*_*j*_ at which *j* was infected:

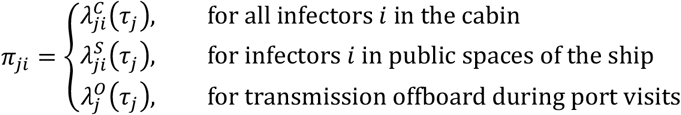

The set of sources of infections *k*_*j*_ represents the probabilistically reconstructed transmission chain for a given set of parameters and imputed infection times.

#### 1.2 Initialization of times of diagnosis and infection

Since for each GI case on the ship only the date of diagnosis is known (measured in days from the cruise start) and not the specific time of diagnosis, we first impute for each case *j* the time of diagnosis *d*_*j*_ by sampling uniformly across the date of diagnosis, excluding night hours (between midnight and 8am).

For each case *j*, we then initialize an imputed time of infection *τ*_*j*_ by sampling values with probability *P*_*I*_(*τ*) of being infected on the time step *τ*, defined as:

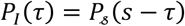

where *P*_*s*_(*x*) is the discretized (on time steps of length 1 hour) and normalized probability distribution of the incubation period, i.e., the probability to develop symptoms after a time *x* since infection. The continuous probability density function of the incubation period used to build *P*_*s*_(*x*) is a log-normal with parameters: *μ*_*s*_ = 0.182 days and *σ*_*s*_ = 0.494 days (mean incubation period: 1.36 days) [S3].

The index case (or cases) that brought the virus on board and started the outbreak must have been infected either before embarkation or during the first port visits. In order to impute the index case(s), we compare the minimum value among the imputed infection times, *τ*_*m*_, with the time of first embarkation on the ship, *T*_*emb*_ (assumed to be at 19: 00 on the first day), and with the time of first port visit, *T*_*f*_. If *τ*_*m*_ < *T*_*emb*_, then the index case was imported at first embarkation on the ship and no further operation is needed. Any other case with time of infection before *T*_*emb*_, besides the one corresponding to *τ*_*m*_, was considered as imported. If *τ*_*m*_ < (*T*_*emb*_ + *T*_*f*_)/2, i.e. the minimum imputed time of infection is closer to first embarkation than to the first port visit, then we still assume that the index case was also imported at first embarkation, and we forcedly reassign the time of infection for the index case as *T*_*emb*_ − 1. All other cases are assumed to be infected during the cruise, including during visits at ports. If *τ*_*m*_ > (*T*_*emb*_ + *T*_*f*_)/2, we forcedly reassign all times of infection comprised between (*T*_*emb*_ + *T*_*m*_)/2 and *T*_*f*_ to *T*_*f*_, under the assumption that they were all index cases infected during the first port visit.

#### 1.3 Calibration

After imputing the times of diagnosis and the starting points for the times of infection of all cases, we estimated the free model parameters (*α, β*_*C*_, *β*_*S*_, *β*_*P*_, *ϕ*), the unknown times of infection *τ*_*j*_ and the source of infection *k*_*j*_ for each case *j*, using a Monte Carlo Markov Chain (MCMC) procedure. The MCMC calibration was then repeated for Z=50 times different model initializations.

The overall likelihood of the observed cases, given the set of parameters *ω* = (*α, β*_*C*_, *β*_*S*_, *β*_*P*_, *ϕ*), the times of infection *τ*_*j*_, and the sources of infection *k*_*j*_, is given by:

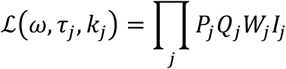

where *P*_*j*_ represents the likelihood that j was infected by *k*_*j*_:

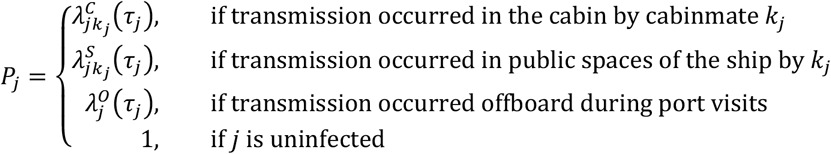

*Q*_*j*_ represents the likelihood that j was not infected:

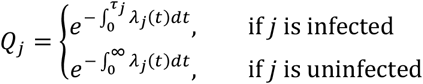

The factor *W*_*j*_ is the contribution to the likelihood of the incubation period for individual *j*, that is *W*_*j*_ = *P*_*s*_(*s*_*j*_ − *τ*_*j*_), where *s*_*j*_ is the known time of symptom onset and *τ*_*j*_ is the inferred time of infection. Finally, *I*_*j*_ = *α* if the individual *j* has been imported from outside before embarkation, and *I*_*j*_ = 1 − *α* if the infection of *j* occurred onboard or if *j* remained uninfected during the whole voyage.

At each step, all parameters (*ω* and infection times *τ*_*j*_) are updated using reversible normal jumps. The MCMC algorithm was run for 400000 iterations, with the first 300000 discarded as burn-in. We discarded (assigning a zero likelihood) all the realizations with infection times that involve a situation with no possibility of transmission (e.g., the infection time of a given individual is sampled during night hours, but there are no other cabinmates infected yet). M=2500 samples were drawn from the posterior distributions obtained by the MCMC.

Results from the *M* × *Z* = 125000 MCMC were pooled together to obtain the final parameter distribution and the distribution of the sources of infection for each case. Figure S1 reports the posterior distributions of estimated parameters.

**Figure S1.**
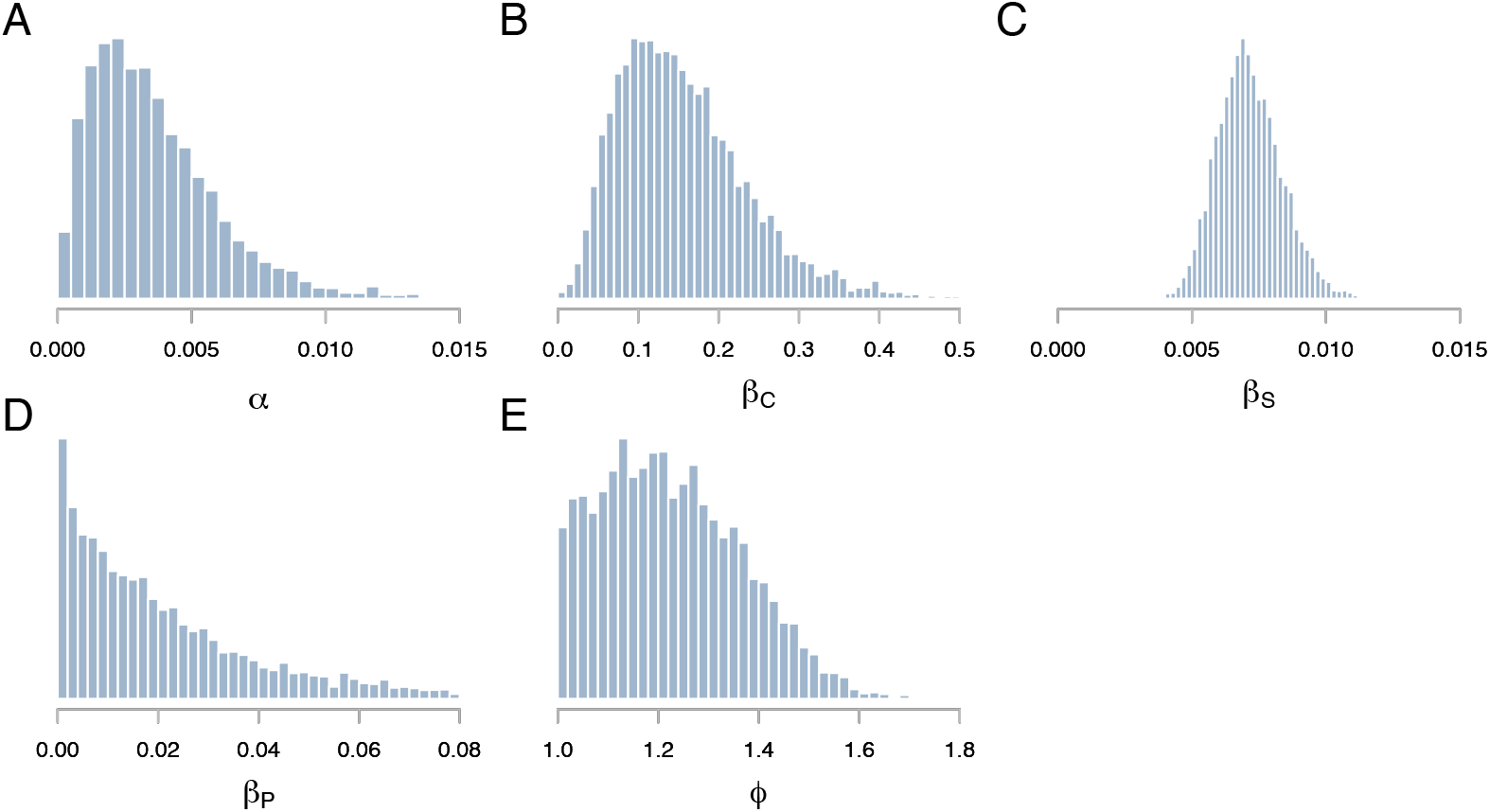
Pooled posterior distributions of free parameters of the model obtained by MCMC. **A** norovirus prevalence in the general population; **B** transmission rate within cabins; **C** transmission rate in public spaces; **D** transmission rate offboard during port visits; **E** increased relative transmissibility in public spaces for cabinmates.

#### 1.4 Sensitivity analyses with underreporting

Available estimates suggest that between 23% and 40% of cases may go undetected in a norovirus outbreak on a cruise ship [S4,S5]. In the baseline analysis, we implicitly assumed that all infections in the outbreak have been reported. In this section we test the robustness of our conclusions when accounting for underreporting. To tackle this question, we apply the same model to a line list of cases where, in addition to reported cases in the data, a number of underreported cases is synthetically generated in such a way that the proportion of underreported cases over the total is 23% or 40%. For each synthetically generated unreported case, the symptom onset date is assigned with multinomial probability based on the observed time series by date of symptom onset. The hour of symtptom onset is assigned with uniform probability throughout the day of symptom onset. We sampled whether the individual had vomiting or diarrhea, with probabilities given by the proportions of cases with vomiting (*p*_*V*_ = 79%) or diarrhea (*p*_*d*_ = 99%) observed during the cruise among reported cases.

The figures below demonstrate that our main conclusions from the baseline analysis remain consistent, even when underreporting is included into the model. Figure S2 shows trends similar to the baseline for both the distribution of secondary cases generated by infectors and the proportion of secondary cases ranked by infectors.

**Figure S2.**
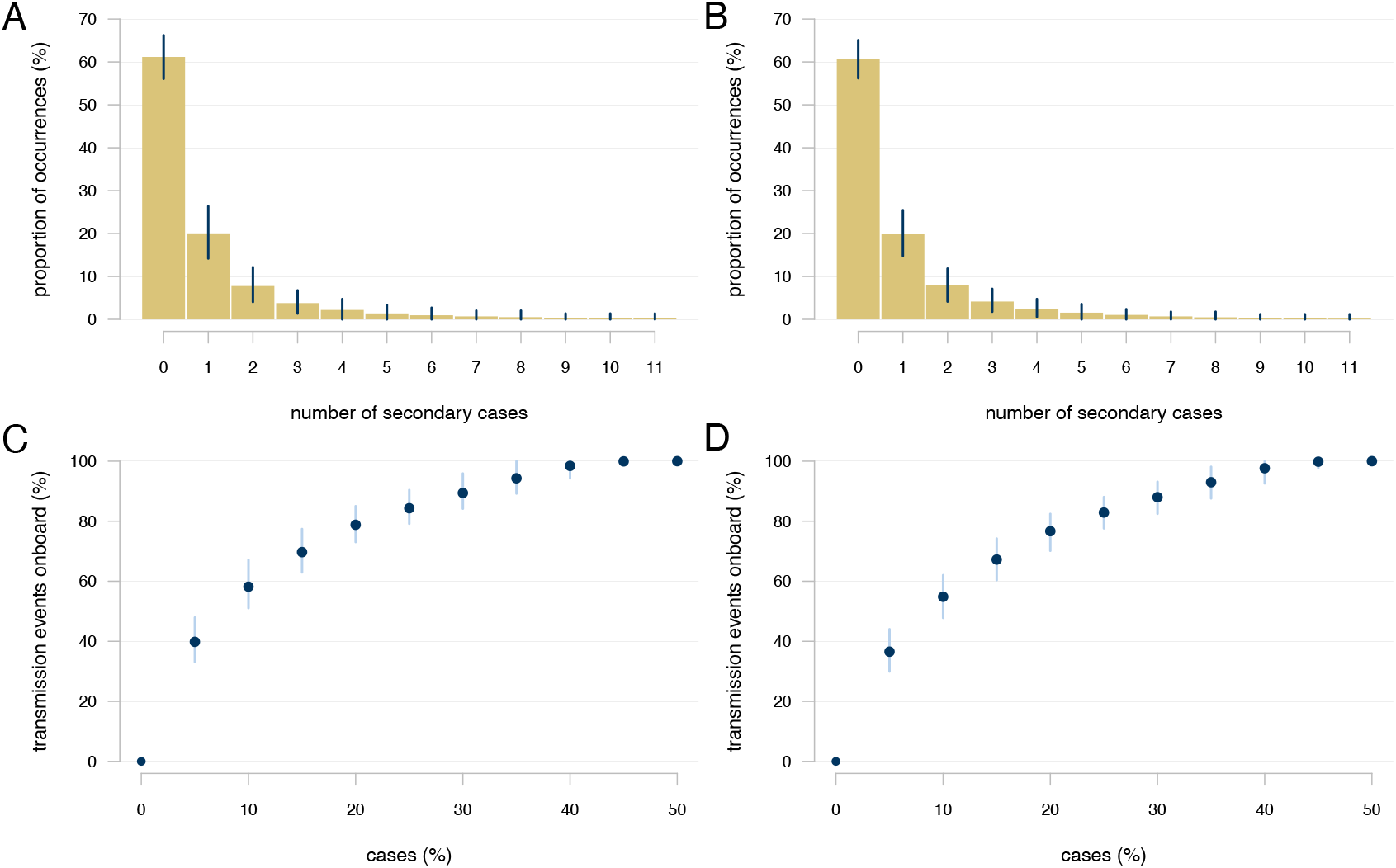
**Top**: Distribution of the number of secondary cases generated by infectors. The bars indicates the average value, while the error bars indicate the 95%CrI over all the reconstructed chains. **A** Underreporting of 23%; **B** Underreporting of 40%; **Bottom**: Cumulative proportion of secondary cases ranked by infectors. The points indicates the average value, while the error bars indicate the 95%CrI over all the reconstructed chains. **C** Underreporting of 23%; **D** Underreporting of 40%.

**Figure S3.**
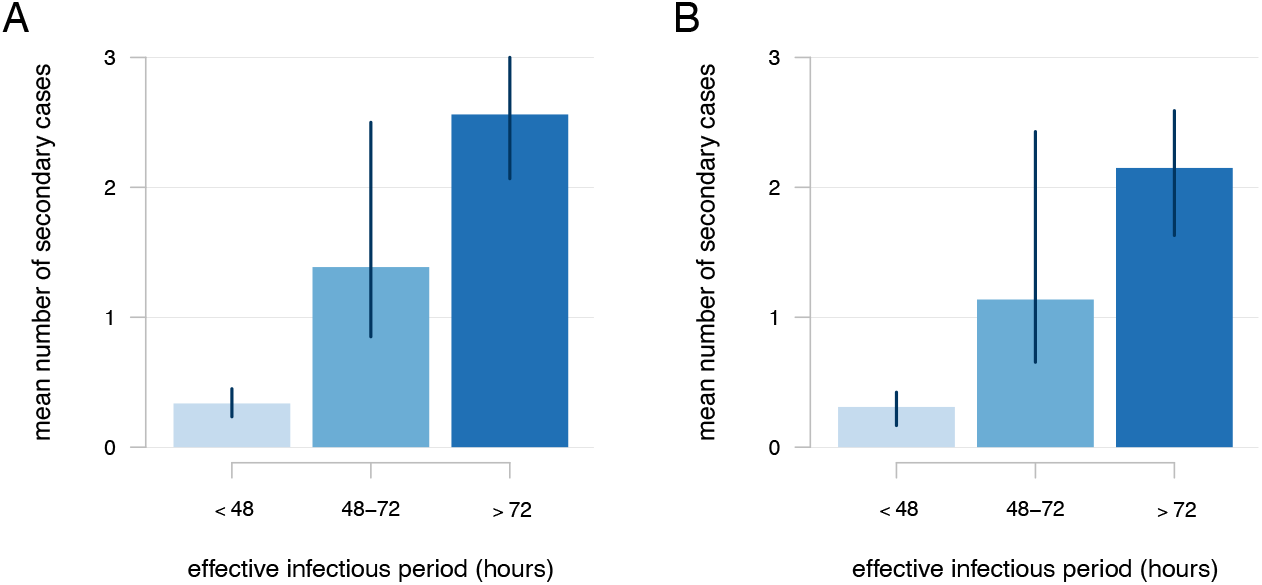
Number of secondary cases stratified by effective infectious period. The bars indicates the average value, while the error bars indicate the 95%CrI over all the reconstructed chains. **A** Underreporting of 23%; **B** Underreporting of 40%.

**Figure S4.**
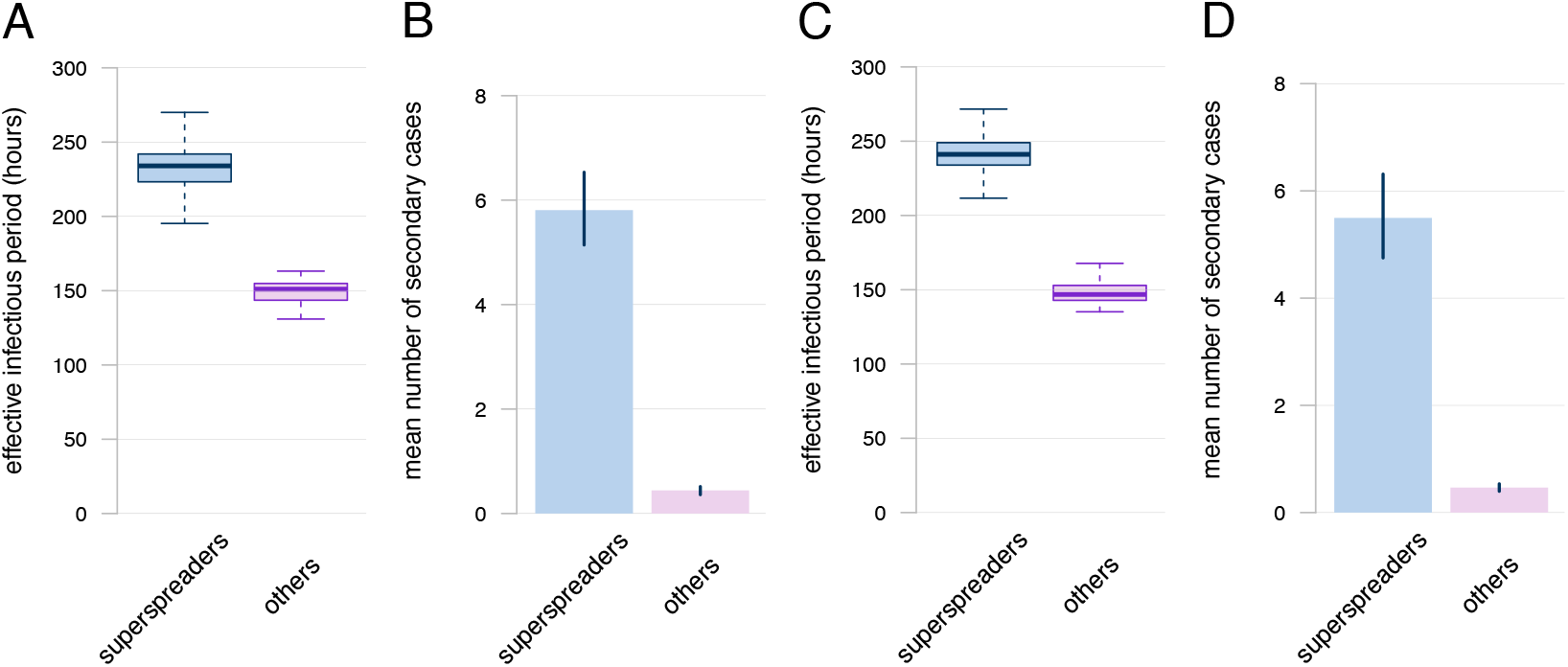
Characteristics of superspreaders compared to other infectors. **A,C** Boxplot of effective infectious period distribution of superspreaders (top 10% of infectors) and of other cases; **B**,**D** Average number of secondary cases generated by superspreaders (top 10% of infectors) and by other cases. The bars indicates the average value, while the error bars indicate the 95%CrI over all the reconstructed chains. **A**,**B** Underreporting of 23%; **C**,**D** Underreporting of 40%. We estimate that the average percentage of undiagnosed superspreaders is 27% (95%CrI: 7% - 47%) when underreporting is 23% and 43% (95%CrI: 23% - 63%) when underreporting is 40%.

We define the “effective infectious period” as the interval during which individuals may transmit the infection. For reported cases, this corresponds to the time between the infection and diagnosis, while for underreported cases it is the time between the infection and the end of the cruise.

Figure S3 shows that the mean number of secondary cases generated by infectors increases with a longer effective infectious period, consistent with findings from the baseline analysis (where, the effective infectious period corresponds entirely with the infection diagnostic delay). Figure S4 illustrates the differences in the effective infectious period and the mean number of secondary cases between superspreaders (top 10% infectors) and other cases. Consistent with results from the baseline analysis, both the effective infectious period and the mean number of secondary cases are significantly higher for superspreaders. Finally, Figure S5 shows, in line with the baseline analysis, a decreasing trend in the distributions of the effective infectious period over time of infection of cases (measured since first embarkation), along with a decaying distribution of the time-varying reproduction number.

**Figure S5.**
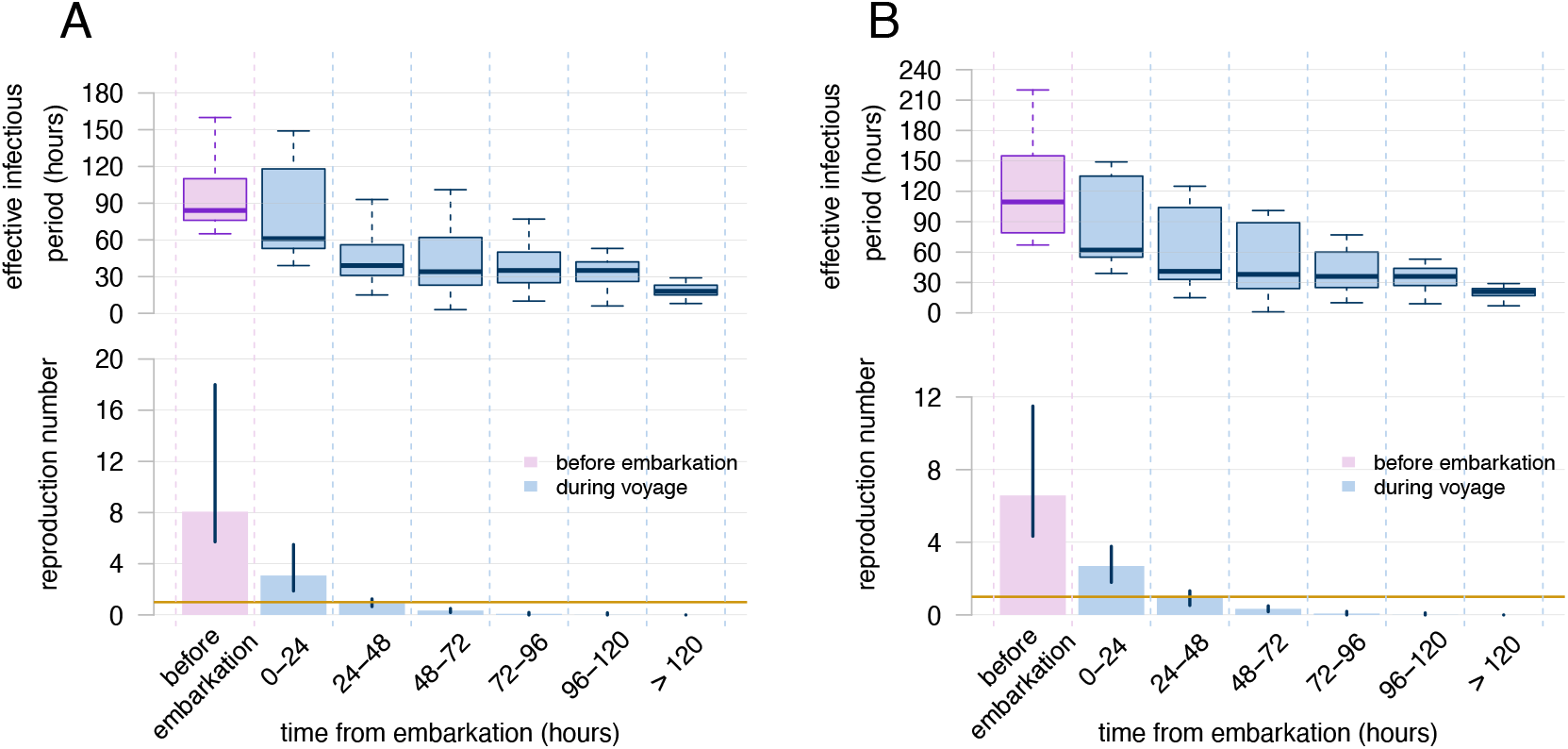
Reproduction number and diagnostic delay over the course of the outbreak. The box plots show the distribution of effective infectious period (in hours) for cases infected each day of the voyage (top). The bar plot (bottom) represents the estimated reproduction number for cases before and during the voyage. The bars indicates the average value, while the error bars indicate the 95%CrI over all the reconstructed chains. **A** Underreporting of 23%; **B** Underreporting of 40%.

### 2 Branching process model

#### 2.1 Model description

To assess the overall effectiveness of alternative isolation protocols and the impact of diagnostic delays, we implemented a stochastic branching process model [S6] that allows to simulate the generation of secondary cases in the considered outbreak under different assumptions.

In the branching process, each infector generates a random number of secondary infections, which is sampled from an offspring distribution. The mean of this distribution corresponds to the basic reproduction number (*R*_0_). Based on the evidence of superspreading that we found via transmission chain reconstruction, we considered a negative-binomial distribution for the offspring distribution, with a mean of *R*_*A*_ and an overdispersion parameter *ν* [S7]. The branching process was applied to the overall population carried on the ship with no distinction between passengers and crew. We assumed homogeneous mixing and disregarded heterogeneities in transmissibility by symptom status.

In the data, two individuals had symptom onset on day 2 of the voyage. We initialized the model with two index cases, assuming these cases were infected before first embarkation. Their infection times were sampled from the same incubation period distribution considered before [S3] and constrained to occur before embarkation (since a negligible number of individuals was infected in port visits, according to the reconstructed chains).

Then, for each newly infected individual *i*_*t*_ (where *i*_*t*_ = 1, …, *I*_*t*_), we sample the number of secondary cases 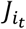 they generate from the negative-binomial offspring distribution:

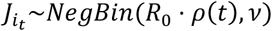

where *ρ*(*t*) represents the proportion of susceptible individuals at time *t*, computed as the difference between the total population size, *N*_*tot*_= *N*_*pax*_ + *N*_*crew*_ = 1229 + 487 = 1716, and the cumulative number of infections before time *t, N*_*I*_(*t*), rescaled by *N*_*tot*_.

For each secondary case 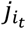 (where 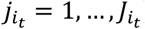), we assign an infection time by sampling from the generation time distribution; infections occurring during the infector’s isolation period, which begins upon diagnosis and lasts for a time that depend on the considered scenario, were discarded and considered as averted by the intervention; those occurring after the end of the cruise were also discarded. As a result, the actual number of realized infections was always lower than the theoretical number of secondary cases sampled from the offspring distribution even in scenarios without isolation. For each infection that was not discarded, we assign a time of symptom onset by adding to the time of infection a sample from the incubation period; and we assign a time of diagnosis by adding to the time of symptom onset a sample from a diagnostic delay distribution which depends on the day of symptom onset of the case (see Section “Empirical diagnostic delay distribution” below) and on the isolation scenario. Secondary infections for which diagnosis occurred after the end of the voyage were considered as unreported but could contribute to transmission on-board.

#### 2.2 Calibration

To calibrate the branching process model to the observed time-series of norovirus cases by symptom onset and diagnosis, we used a combination of grid search and particle filtering [S8]. The two free model parameters were the basic reproduction number *R*_0_ and the overdispersion of the offspring distribution *ν*.

For particle filtering, we used a discretized version of the branching process model described above, with a time step of Δ*t* = 1 hour. For a given parameter set, the performance of the model was evaluated at each time step using a mean-squared error (MSE) function, comparing the number of reported cases in simulated trajectories against the observed time series, aggregated by time of symptom onset over 4-hour intervals and by date of diagnosis over 24-hour intervals. At each aggregated time of analysis (i.e., 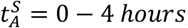, 4 − 8 *hours*, 8 − 12 *hours*, …, for trajectories by time of symptom onset and 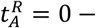, 24 − 48 *hours*, 48 − 72 *hours*, …, for trajectories by date of diagnosis), the MSE of a particle *w* is defined by:

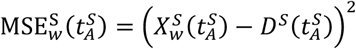

and

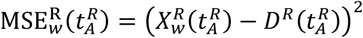

where 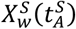 is the trajectory of particle *w* by time of symptom onset; 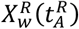 is the trajectory of the particle *w* by date of diagnosis; 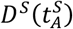 is the time series of observed cases by time of symptom onset; 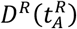 is the time series of observed cases by date of diagnosis; The resulting total score of particle *w* was given by 1/MSE_*w*_, where:

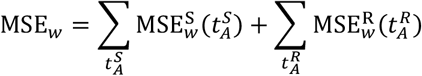

The particle filtering algorithm consists of two steps:

1. At each time step *t*, W=1000 particles were generated and the M=100 with the best scores (i.e., the 100 lowest MSE_*w*_) were selected;
2. From the top M particles, W particles were sampled with replacement, with probabilities weighted by their score. These W particles were then used to reinitialize the model to generate the next time step (*t* + 1).

The frequent resampling of particle filtering can lead to the so-called degeneracy problem, where only a few particles dominate the simulation, reducing the variability necessary to accurately represent the outbreak dynamics [S8]. To prevent this and ensure sufficient diversity among the particles, we repeated the particle filtering process S=5 times, each time starting from different random seeds.

The selected M particles from all S procedures were pooled together and the final score for the given parameter set was given by their average mean squared error:

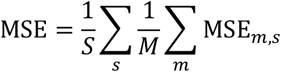

To estimate parameter values, we evaluated the MSE for values of *R*_0_ between 8 and 24, with steps equal to 0.25, and values of *ν* from 0.1 to 0.6, with steps equal to 0.02, using a grid search (i.e., testing all combination of parameter values, corresponding to a total of 65×26=1690 parameter sets explored). Figure S6 illustrates the values of the MSE across the explored parameter sets. Considering the *K*=100 parameter sets with lowest MSE, the estimated average basic reproduction number is 15.6 (95%CrI 11.5-20.4) and the estimated average overdispersion is 0.13 (95%CrI 0.10-0.18). Figure S7 shows the fit of the *M* × *S* × *K* trajectories against observed data.

**Figure S6.**
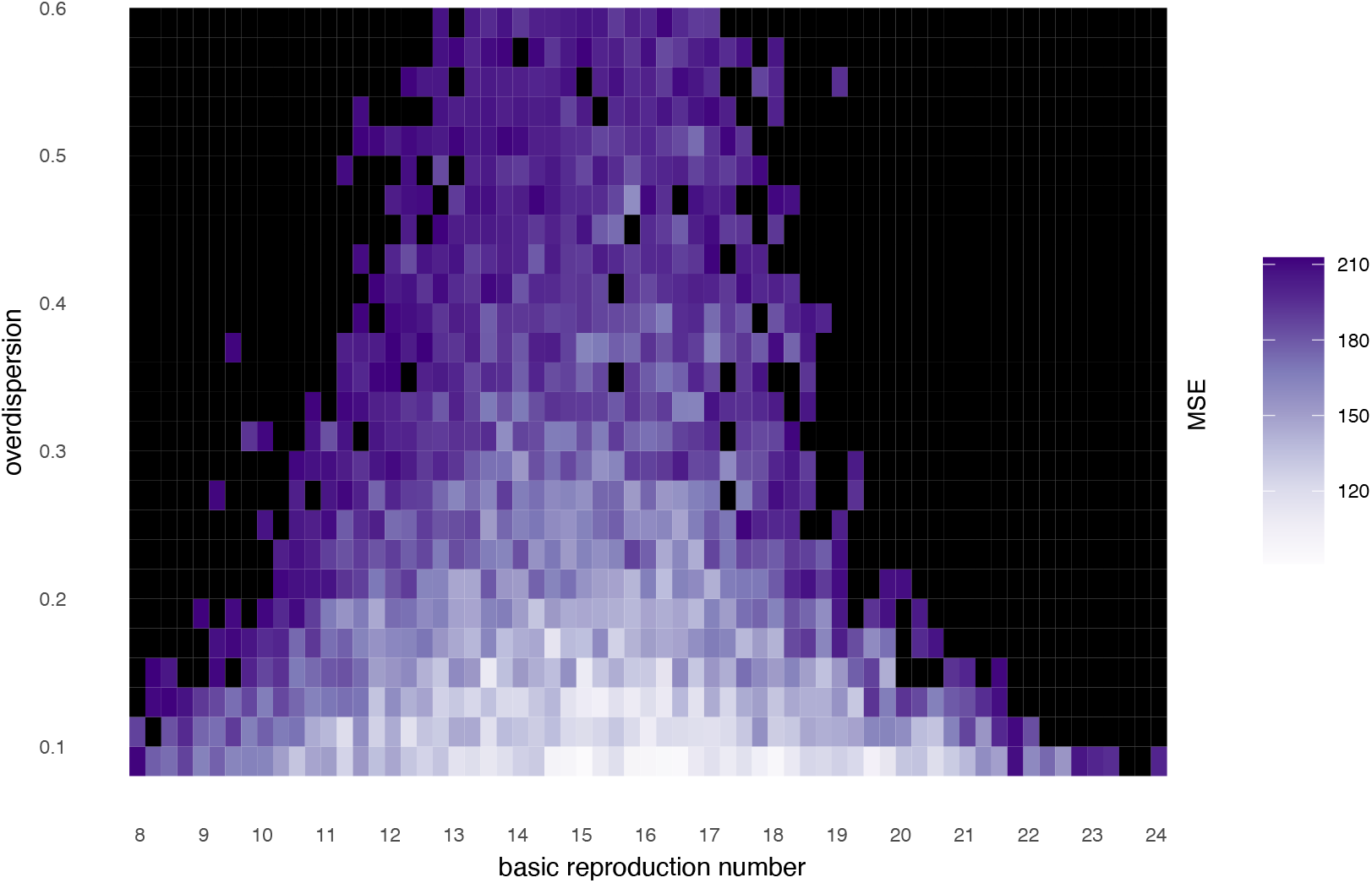
Grid of mean squared error (MSE) values across parameter sets for basic reproduction number and overdispersion. Lower MSE values indicate better fits of the model to observed data.

**Figure S7.**
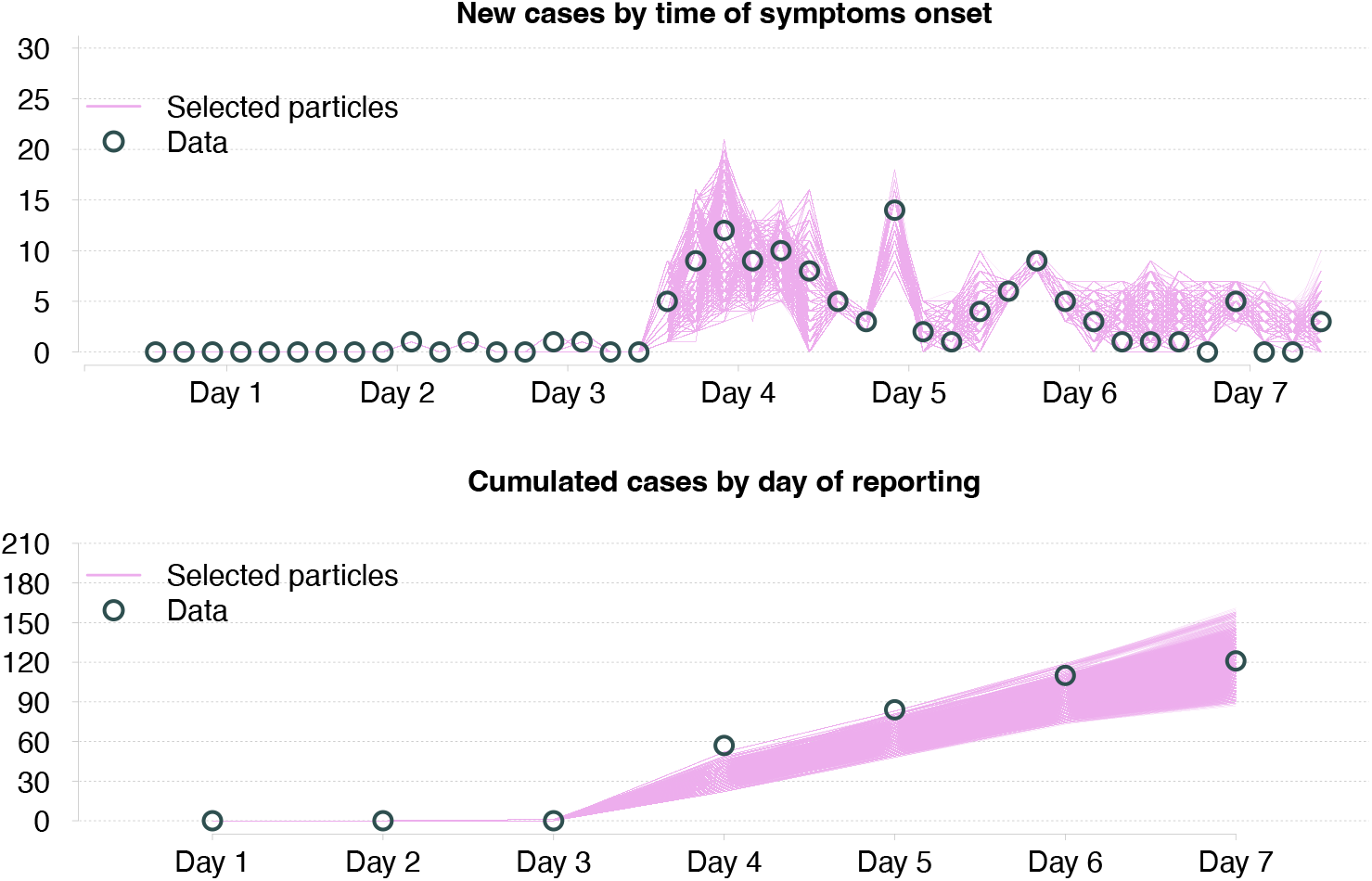
Comparison of pooled selected particles from the branching process model for the top 100 parameter sets with actual data for the time series. **Top**: New cases by time of symptom onset; **Bottom**: Cumulative cases by reporting day (day of diagnosis).

#### 2.3 Empirical diagnostic delay distributions

The diagnostic delay from symptom onset refers to the interval between the onset of symptoms and the diagnosis of cases. Since the available data only included the day of diagnosis, not the specific time, the probability distribution of the diagnostic delay was estimated by imputing for all cases a diagnostic time sampled uniformly over the day, under the constraint that diagnoses occurred during daytime (between 08:00 and 23:59) and always after the onset of symptoms (since no norovirus screening was implemented on the ship). The imputation process was repeated N_A_=50 times, and a distribution of diagnostic delays was obtained by pooling together diagnostic delays from the N_A_ imputations. Because cases with later symptom onset tended to have smaller diagnostic delays, due to the right censoring imposed by the end of the cruise, we estimated different diagnostic delay distributions based on the day of symptom onset (Figure S8).

**Figure S8.**
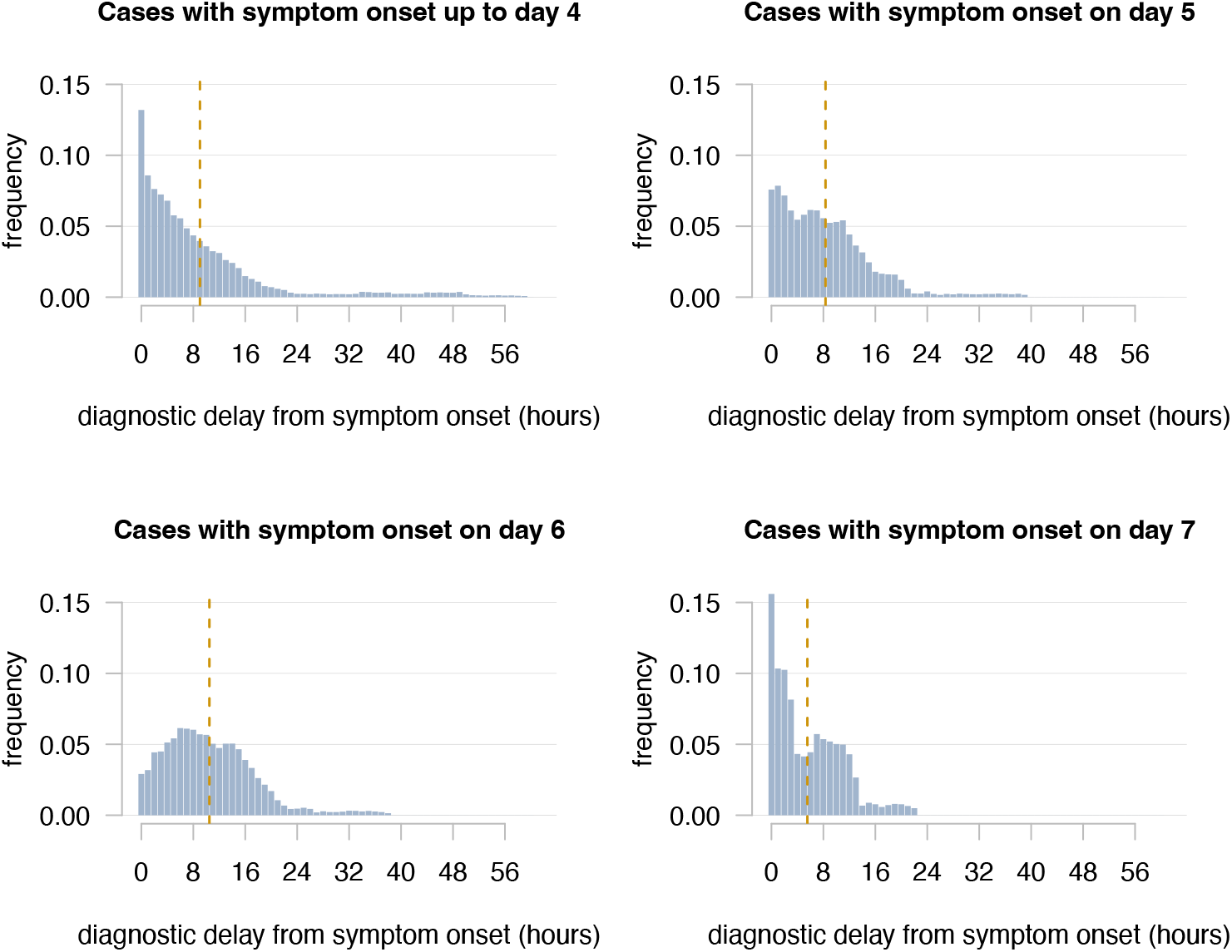
Empirical diagnostic delay distribution, disaggregated by day of symptom onset. Vertical dashed lines indicate mean values of distributions.

#### 2.4 Output analysis

A schematic description of scenarios considered is presented in Table S1. Results for all scenarios were obtained by running L=100 stochastic iterations with the top *K* parameter sets.

**Table S1.** Synopsis of scenarios.

| Name | Description | Duration of isolation | Diagnostic delay distribution |
| --- | --- | --- | --- |
| S<sub>0</sub> | baseline | 72h | empirical |
| S<sub>1</sub> | no isolation | 0h | empirical |
| S<sub>2</sub> | 24h isolation | 24h | empirical |
| S<sub>3</sub> | 48h isolation | 48h | empirical |
| S<sub>4</sub> | baseline isolation, increased diagnostic delay | 72h | increased by 50% |
| S<sub>5</sub> | baseline isolation, reduced diagnostic delay | 72h | reduced by 50% |
| S<sub>6</sub> | perfect isolation | until end of cruise | no delay (immediate diagnosis upon symptoms) |

We computed three main outputs for each scenario: the effective reproduction number, the relative change in the total number of cases observed during the cruise, and the probability of an outbreak.

The effective reproduction number was computed by averaging the mean number of secondary infections caused by cases that were either imported before embarkation or infected within the first two days of the voyage. This choice is based on the observation that the number of secondary cases tends to decline sharply for infections occurring later in the cruise due to the right truncation caused by the end of the cruise (see Figure S9 for the reproduction number disaggregated by time of infection of the cases).

**Figure S9.**
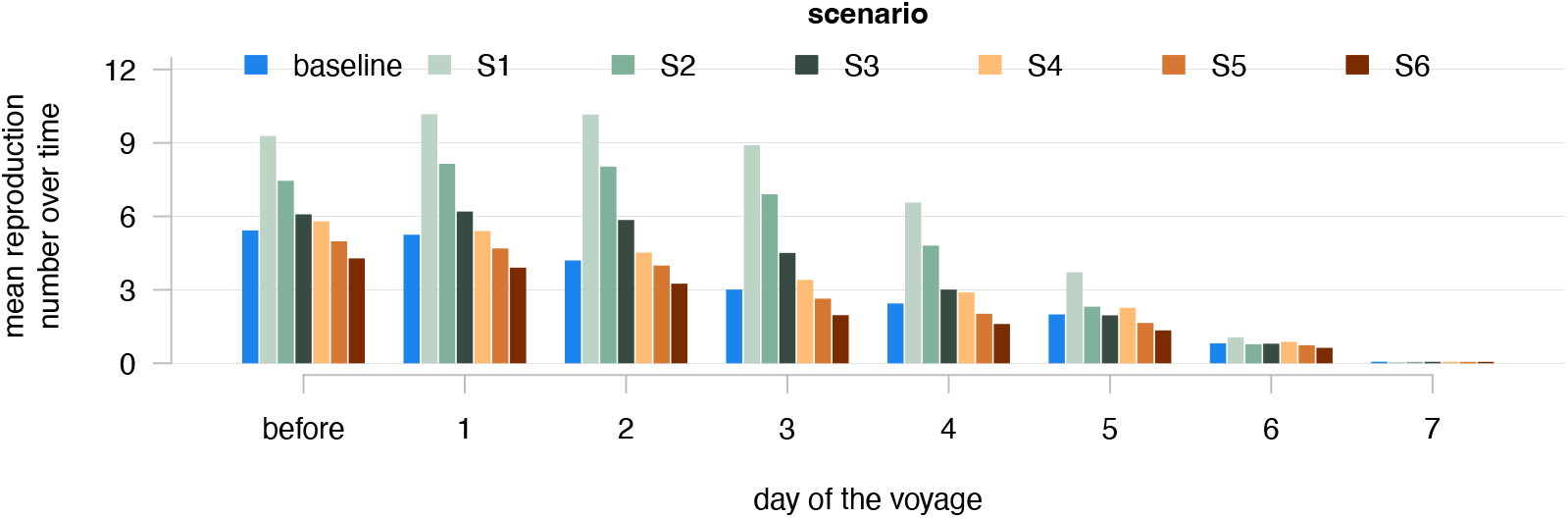
Mean time-dependent reproduction number R_t_ across alternative scenarios. Bars indicate mean values over 100 simulations.

For each scenario *X*, the relative change Δ_*X*_ in the total number of cases was calculated as the percentage change in the mean number of cases expected for the considered scenario, 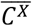 (calculated over the *L* × *K* simulations), compared to the mean number of cases 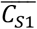 expected for scenario S1, where no isolation was implemented:

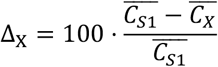

where 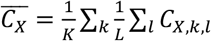 and *C*_*X, C, l*_ represents the number of cases generated in the *l-*th simulation with the *k*-th parameter set in scenario *X*.

The probability of an outbreak was determined as the proportion of the *L* × *K* simulations where the number of reported cases was larger than 2% of the total passenger population, corresponding to a threshold outbreak size of *S*_*thr*_ = 0.02 ⋅ *N*_*pax*_= 25 cases. The threshold was referred only to the passengers population as these represented the vast majority of reported cases in the considered data.

## Notes

### Competing Interest Statement

The authors have declared no competing interest.

### Funding Statement

This study was conducted in the framework of the Healthy Sailing project which received funding from the European Union's Horizon Framework Programme under grant agreement no. 101069764.

### Author Declarations

Ethics Committee of University of Thessaly gave ethical approval for this work

